# Impact of the Novel Coronavirus Disease (COVID-19) on Treatment Adherence and Sleep Duration in Obstructive Sleep Apnea Patients Treated with Positive Airway Pressure

**DOI:** 10.1101/2020.06.28.20141994

**Authors:** Salma Batool-Anwar, Olabimpe S. Omobomi, Stuart F. Quan

## Abstract

**Objective:** To examine the effect of COVID-19 on treatment adherence and self-reported sleep duration among patients with Obstructive Sleep Apnea (OSA) treated with positive airway pressure (PAP) therapy.

**Methods:** Retrospective review of medical records of patients seen in Sleep and Circadian Clinic at Brigham Health during the immediate period of one month after the national lockdown was announced on March 15, 2020. Patients with OSA were included only if PAP adherence data was available in the 12-months prior and in the month after the lockdown. Patients with other sleep disorders and OSA patients without the adherence data were excluded.

**Results:** Mean age was 63.5± 13.9 years, 55% of the participants were men, and mean BMI was 31.8 ± 7.9 kg/m^2^. Severe OSA was noted among 59.5% compared to 29.3% moderate, and 11.2% mild OSA. Increased number of patients reported insomnia after the lockdown (41% vs 48%, *p= 0*.*02)*. Gender stratification noted worsening insomnia only among women. There was no significant difference in PAP adherence as measured by the hours of use, self-reported sleep duration or in the use of sleep medications.

**Conclusion:** Post COVID-19 lockdown had a negative impact on sleep as evidenced by increased reporting of insomnia particularly among women, but no impact on PAP adherence or self-reported sleep duration.

## Introduction

In December 2019, a severe and lethal respiratory infection appeared in Wuhan, China and subsequently was found to be caused by a novel coronavirus, SARS-CoV-2.^1^ The worldwide pandemic of COVID-19 rapidly ensued. The first case of COVID-19 in the United States occurred in January 2020 ^2^ and quickly spread across the country. This led on March 15, 2020 to a voluntary national shut down with closure of schools and businesses. As a result, most of the United States population confined themselves to their residence either unemployed or working from home.

A well-functioning sleep-wake cycle is vital to our health, immune system and prevention of chronic diseases. At least 7 hours of sleep are recommended for optimal health by the American Academy of Sleep Medicine.^3^ During reports of previous natural disasters and other crises where the general population was under severe psychological stress, sleep disturbances, particularly insomnia, increased.^4-6^ Moreover, during the 2003 SARS outbreak, sleep difficulties were common among family members of patients.^7^ Similarly, initial reports from China during the current COVID-19 pandemic have documented increased rates of insomnia and anxiety.^8^ However, despite the relative frequency of such disasters and crises, there are few reports of their impact on sleep duration. A review of disaster medical records after the 2011 Great East Japan earthquake noted an increase in sleep deprivation especially among women, but hours of sleep were not reported.^5^

Obstructive sleep apnea (OSA) may affect as many as 26% of adults.^9^ Positive airway pressure (PAP) is the most common form of treatment and improves the poor sleep quality and daytime sleepiness associated with OSA. However, PAP adherence is sometimes difficult to achieve; co-morbid insomnia and anxiety maybe factors that are important in limiting usage.^10^ In this study, we hypothesized that hours of objective PAP adherence and self-reported sleep duration would be adversely affected by the stress and anxiety caused by COVID-19 and home confinement.

## Methods

### Design

A retrospective review of medical records was conducted of patients who attended the Sleep and Circadian Disorder clinics at Brigham Health before and after the announcement of the COVID-19 lockdown. The study was approved by the Institutional Review Board at Brigham Health.

### Study population

The study population consisted of patients with OSA seen in the clinic during the immediate period of one month after the national lockdown was announced on March 15, 2020. A total of 588 clinic encounters were reviewed of which 30 patients had in person clinic visits, and 558 were done virtually. Patients with OSA were included in this study if CPAP adherence data was available in the 12 months prior and in the month after the COVID-19 lockdown. Patients with other sleep disorders such as restless legs syndrome, narcolepsy/idiopathic hypersomnolence, and insomnia without OSA were excluded. Additionally, patients with OSA on alternative treatments were also excluded. This resulted in 123 patients with data from both time periods who were included in the analytic cohort.

### Data Collected

Using the electronic medical records (EMR), information on age, gender and body mass index (BMI, kg/m2) was collected. Using clinician documentation, we collected data on OSA severity, self-reported sleep duration, subjective complaints of insomnia and anxiety, and the use of sedative-hypnotic medications. Mean hours of positive airway pressure (PAP) usage was obtained from the 30 day compliance report downloaded from the PAP device which had been scanned in patient’s EMR.

### Statistics

All statistical analyses were done using STATA version 11 (StataCorp, LLC, College Station, TX, USA). For baseline characteristics, mean (SD) for continuous variables and number and percentages for categorical variables were calculated. To test for differences in variables before and after the CoVID-19 imposed lockdown Student’s paired t-test was used for continuous variables (self-reported sleep duration and objective PAP use) and McNemar’s chi-square test was employed for binary paired data (anxiety, insomnia, and use of medications). Statistical significance was set at a p value <0.05, two-tailed.

## Results

Mean age of the cohort was 63.5± 13.9 years; over half were men (55%). Mean BMI was 31.8 ± 7.9; mild, moderate and severe OSA were present in 11.2, 29.3 and 59.5% respectively.

As shown in the Table, there were no differences in hours of PAP use or self-reported sleep duration. Stratification by OSA severity did not materially change the results (data not shown). Data were available for the presence of insomnia, anxiety and sedative-hypnotic medications in 98, 73 and 98 patients respectively. After the lockdown, there was an increase in the number of patients with insomnia (41 vs. 48%, p=0.02). Further stratification by gender demonstrated worsening insomnia only among women (data not shown). There also was no difference in the use of sleep medications post COVID-19.

**Table.**
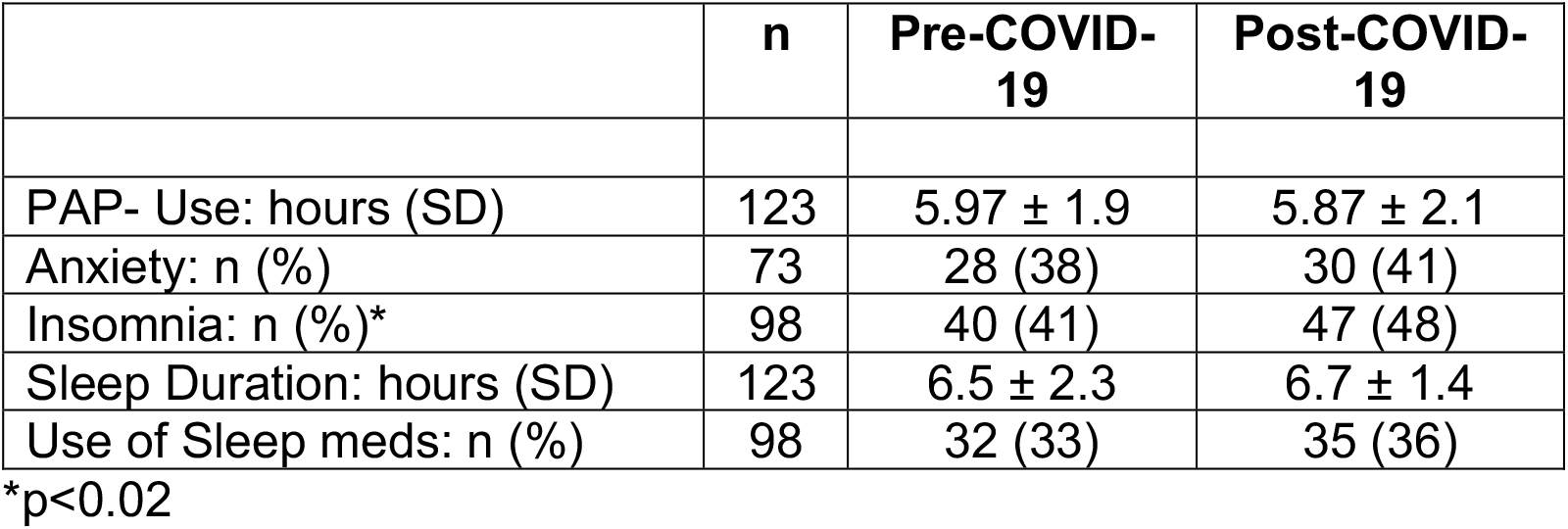
Effects of COVID-19 Lockdown.

## Discussion

Global crises including pandemics, natural disasters and wars are a known source of psychological stress and may exacerbate anxiety and insomnia, and adversely impact sleep quality in vulnerable populations.^4-6^ Furthermore, sleep deficiency is common among American adults. According to the Centers for Disease Control and Prevention, 35% report less than the recommended 7 hours of sleep per night (https://www.cdc.gov/sleep/data_statistics.html). Although the presence of the lockdown and home confinement provided the opportunity to increase sleep duration, we did not observe any change in this small cohort. This is similar to reports of unchanged sleep duration among Israelis during Scud missile attacks on nights without missile attacks ^6^. However, it stands in contrast to a prior study which reported evidence of sleep deprivation after an earthquake.^5^

We also did not find a significant difference in the recorded hours of PAP usage before and after the lockdown. This is in contrast to the findings of a recent study that reported an increase in PAP usage in France after their lockdown.^11^ Importantly for clinicians, however, it appears that living in an area with a large number of COVID-19 cases does not decrease PAP adherence.

Even though subjective and objective sleep duration were not different before and after the lockdown, we noted a higher prevalence of insomnia in the month after the lockdown was announced. This indicates that the patients in this cohort had an insomnia phenotype without objective short sleep duration. Insomnia is a known consequence of acute stress and is often mediated via anxiety related to ongoing stressful life events. Insomnia may present as difficulty with initiating or maintaining sleep with or without short sleep duration or perceived poor sleep quality.^12^ Prior studies have documented increased insomnia symptoms during times of crises such as war ^13^, epidemics^14^ and natural disasters.^15^ Thus our finding is consistent with a recent study also performed during the current pandemic.^16^ Gender stratification showed that the observed increase in insomnia symptoms after the lockdown occurred in women. This is similar to findings of a study done during the Gulf war ^6^ and consistent with the known higher prevalence in women.^17^

Anxiety is commonly comorbid with insomnia,^18^ but we did no observe any change in prevalence after the lockdown. However, our study did not examine whether there was any exacerbation of chronic anxiety symptoms after the lockdown in those who were already afflicted.

To our knowledge this is the first study examining the effect of the COVID-19 related lockdown on self-reported sleep duration for patients using PAP. However, we acknowledge some limitations. First, we reviewed medical records of patients seen in the sleep clinic and did not have a healthy control group. However, our primary goal was to identify changes in sleep duration and hours of PAP usage after the onset of the pandemic. A second limitation is the short duration of the study and the limited cohort size as only records of patients seen immediately after the lockdown were reviewed. It is possible that psychosocial stress and worsening mental health in the face of lockdown and economic downturn prevented some individuals from seeking medical help.

In conclusion, the post COVID-19 lockdown has negatively affected sleep resulting in increase in insomnia without changes in PAP usage and self-reported sleep duration. Future long-term studies are needed to assess the effect of SARS-COV2 related stress on sleep duration and hours of PAP usage.

## Data Availability

Data are available from the authors

